# Cost-effectiveness of alternative cascade screening strategies for familial hypercholesterolemia with realistic cascade screening acceptance rates and use of high-cost drugs

**DOI:** 10.1101/2025.01.17.25320710

**Authors:** Jing Lou, Kok Joon Chong, Sharon Li Ting Pek, Yasmin Bylstra, Chester Lee Drum, Weng Khong Lim, Yi Wang, Khung Keong Yeo, Subramaniam Tavintharan, Hwee-Lin Wee

**Affiliations:** Saw Swee Hock School of Public Health, National University of Singapore; Khoo Teck Puat Hospital, Singapore; SingHealth Duke-NUS Institute of Precision Medicine, Singapore; National University Hospital, Singapore; SingHealth Duke-NUS Genomic Medicine Centre, Singapore; Cancer & Stem Cell Biology Program, Duke-NUS Medical School, Singapore; Laboratory of Genome Variation Analytics, Genome Institute of Singapore; National Heart Centre Singapore, Singapore; Department of Pharmacy, Faculty of Science, National University of Singapore

**Keywords:** familial hypercholesterolemia, cascade screening, cascade acceptance rates, PCSK9 inhibitor

## Abstract

**Background:** Cascade screening (CS) for familial hypercholesterolemia (FH) is cost-effective in many countries. However, most existing economic evaluation studies (i) ignored or overstated first-degree relative (FDR) participation rate (as 60-100%), (ii) did not consider novel and expensive therapies, e.g. PCSK9 inhibitors (PCSK9i), (iii) were conducted outside of Asia. In Singapore, FDR participation rate is about 25% among probands who have known pathogenic variants.

**Aims:** To evaluate and identify drivers of cost-effectiveness of CS protocols for FH

**Methods:** Four CS protocols, which vary in the application of genetic testing, were examined using a hybrid decision tree-Markov model. Sensitivity analysis and scenario analyses were conducted to identify drivers of cost-effectiveness.

**Results:** All CS protocols are likely to be cost-effective (probabilities of being cost-effective: 85%-95% when no access to PCSK9i; 73%-99% when PCSK9i are provided). The most cost-effective protocol differs depending on whether PCSK9i are provided. Cascade acceptance rates are key drivers of cost-effectiveness. Other drivers include timeliness of starting treatment post-screening, age of proband, health-related quality of life loss with cardiovascular disease, prevalence of FH mutation among probands, conventional treatment effect and cost of PCSK9i.

**Conclusion:** CS for FH is cost-effective across most scenarios and assumptions. For better cost-effectiveness, health systems need to look for ways to improve proband’s willingness to share contact of their relatives and relatives’ willingness to be screened. Other ways to improve cost-effectiveness include to select age groups for proband screening, improve screening detection rate among probands, and start timely treatment post-screening.

## 1. Introduction

Familial Hypercholesterolemia (FH) is predominantly an autosomal dominant genetic condition. Carriers have a 50% chance of passing causative genetic variants associated with FH to their offspring(s). Untreated individuals with FH can die from acute myocardial infarction (AMI) at a relatively young age, and for most, this will occur during their prime economically productive age, resulting in significant economic losses to families at the micro level and society at the macro level [1]. Families with FH are likely to inherit lower social-economic status due to cardiovascular events repeated over generations, causing intergenerational immobility. Hence, identifying FH early has the potential to reduce cost to individuals, families and society [2].

Due to the strong genetic linkage, cascade screening, which involves testing family members at risk of FH for the same causative genetic variant and/or elevated LDL-c, has been found to be cost-effective in many countries [3–5]. However, most existing economic evaluation studies (i) ignored or overestimated FDR participation rate for cascade screening (60-100%) and (ii) did not consider novel and expensive therapies, e.g. proprotein convertase subtilisin kexin type 9 inhibitors (PCSK9i). Furthermore, there is no published cost-effectiveness analyses of FH screening from Asia [3,5].

Since 2015, local public hospitals in Singapore have been running a pilot cascade screening program, Familial Hypercholesterolemia: Case Identification, Assessment and Reduction in Adverse Events (FHCARE) [6], with Dutch Lipid Clinical Network (DLCN) criteria, lipid and genetic tests. The acceptance rates for cascade screening were observed to increase over year. As of 2024, among probands detected with pathogenic variants, about 50% of them shared the contact of their first-degree relatives (FDR) and about 50% of the contacted FDR accepted screening, resulting in FDR participation rate at about 25% (50%×50%) among probands (definition of terminologies in **Appendix 1**). Since December 2023, patients in local public hospitals who are diagnosed with FH or cardiovascular disease (CVD) can have access to PCSK9i with subsidized price if LDL-c remains above goal despite maximally tolerated oral lipid lowering agents [7].

This study was undertaken during 2020-2024 to inform the design of FH screening in Singapore, by comparing the cost-effectiveness of various cascade screening protocols that differ in whether genetic test is used and how probands are filtered for the family’s eligibility of cascade screening (whether DLCN is used to filter for probands with higher clinical risk, and whether MLPA is used as complementary test for NGS). We aimed to evaluate cost-effectiveness of the alternative cascade screening protocols, understand the roles of cascade acceptance rates and high-cost drugs, and look for directions to improve cost-effectiveness of cascade screening for FH.

## 2. Methods

### 2.1. Study population

The population examined in this study is any Singapore resident whose LDL-C ≥ 4.9mmol/L and their relatives. Patients with hypercholesterolemia were classified into three groups based on genetic evidence and DLCN criteria (**Appendix 2**):

A. genetic FH: FH patients with causative genetic variants from any of the 8 genes including LDL-R, ApoB, PCSK9, LDL-RAP1, ApoE, ABCG5, ABCG8 and LIPA.
B. clinically FH: DLCN > 5, but without causative genetic variant
C. non-FH: LDL-C above 4.1mmol/L, but DLCN ≤ 5 and without causative genetic variant (The threshold of starting treatment without FH mutation is 4.1mmol/L.)

### 2.2. Setting and location

The study is set in Singapore, an island city-state, whose healthcare system ranks among the best globally. In 2000, the World Health Organization ranks Singapore as the 6^th^ best healthcare system globally while the 2023 Legatum Prosperity Index for “health component” ranked Singapore as the best [8,9]. Nonetheless, while Singaporean residents are encouraged to check their lipid profiles triennially after 40 years old, FH is rarely considered and there formal screening for FH.

### 2.3. Interventions and comparator

Four protocols for identifying probands with FH were evaluated as candidate interventions. They are numbered in the order of increasing use of genetic test. The process diagrams of all screening protocols are in **Appendix 3**.

- Protocol 1 (**Figure A1**) identifies probands based on DLCN criteria only, without genetic test, followed by cascade screening. Individuals whose LDL-c > 4.9mmol/L from opportunistic lipid tests will be referred for screening. If they show up for the screening, they will be scored based on DLCN diagnostic criteria. If a proband has DLCN > 5, the screening coordinator will request the proband to share contact of his/her first-degree relatives (FDR). The coordinator will then invite the FDR for cascade screening. FDR who accept the cascade screening will be screened with lipid tests only. If an FDR’s LDL-c > 4.9mmol/L, the coordinator will request the FDR to share contact of his/her FDR (i.e. second-degree relatives (SDR) of the proband). And the same process of FDR will apply to SDR if their contacts are available.
- Protocol 2 (**Figure A2**) involves NGS offered to probands whose DLCN > 5 followed by cascade screening. If a causative variant is detected by NGS, the screening coordinator will request the proband to share contact of his/her FDR. FDR who accepted cascade screening will be screened by targeted Sanger Sequencing (SS) and lipid test. Only if an FDR has causative variants will the screening coordinator try to reach out to SDR.
- Protocol 3 (**Figure A3**) involves NGS offered to ALL probands followed by cascade screening. Protocol 2 is based on the recommendations from guidelines established in settings with low FH prevalence [10,11], while Protocol 3 is based on guidelines from European Atherosclerosis Society [12] and the National Institute for Health and Care Excellence (NICE) [13] as they have pioneered FH cascade screening programs since 2000s.
- Protocol 4 (**Figure A4**) is an extension of Protocol 3 where MLPA is performed for probands where no causative variants have been detected by NGS, as MLPA is the gold standard for detecting whole-exon copy number variants (CNVs). This complementary step has been considered by some to be essential because around 10% of FH cases are attributed to CNVs [14].

All intervention protocols were compared against status quo in Singapore. Status quo refers to no DLCN, no genetic tests and no cascade screening. We assumed that individuals with LDL-c > 4.9mmol/L are not assessed further for FH and their relatives are not screened. While probands are treated, their relatives who are not on pre-existing treatment will remain untreated until CVD onset. Among probands detected with pathogenic variants and shared contact of their FDR, 50% of the contacted FDR accept screening while this figure is likely to drop to 10% when pathogenic variant is absent or undetected among probands, according to residing experts in FHCARE.

In sensitivity and scenario analyses, we attempt to answer questions that are of relevance of policy makers. (1) how will the provision of high-cost treatments such as PCSK9i affect the cost-effectiveness of FH screening? (2) how will cascade acceptance rates affect the cost-effectiveness of FH screening? (3) what other factors affect cost-effectiveness of cascade screening?

### 2.4. Scenarios without vs with access to PCSK9i

In the base case, PCSK9i is not provided. Hence, treatments for probands or pre-treated relatives are the same across all four screening protocols (P1 to P4). The screening changes the clinical pathway of untreated relatives with causative variants and/or elevated LDL-c by starting treatment for them.

In the scenario analysis where PCKS9i is accessible for patients with FH causative variants, DLCN > 5 or CVD, Protocols P1 and P2 provide PCSK9i to probands whose DLCN > 5 while Protocols P3 and P4 provide PCSK9i to only probands with causative variants.

For relatives, Protocol P1 does not provide PCSK9i to relatives while Protocols P2, P3 and P4 provide PCSK9i to any relatives with causative variants in addition to starting treatment for untreated relatives with mutation and/or elevated LDL-c.

### 2.5. Study perspective

The health system perspective was adopted in this study as per Singapore’s health technology assessment (HTA) agency’s guidelines [15].

### 2.6. Time horizon

Time horizon was assumed to be 100 years to capture full life-time costs and effects regardless of patients’ age, since CVD is a chronic medical condition.

### 2.7. Discount rate

The discount rate was 3% for both costs and health outcomes, as per Singapore’s HTA agency’s guidelines [15].

### 2.8. Description of model

The cost-effectiveness of different proband screening followed by cascade screening protocols was estimated using a hybrid decision tree-Markov cohort model, with the decision tree to reflect the once-off screening process and the Markov cohort model to reflect patients’ follow-up pathway after screening.

The decision tree consisted of a main tree for proband screening (**Figure A5** in **Appendix 4**) and two levels of subtrees for cascade screening of FDR and SDR (**Figure A6** in **Appendix 4**). The main tree is attached to a Markov model, that reflects the proband’s follow-up pathway, and the first subtree, for cascade screening of FDR (FDR subtree). Probands picked up by opportunistic lipid tests were assumed to be previously undiagnosed with FH. The FDR subtree is attached to a Markov model that reflects the FDR’s follow-up pathway and a second subtree for cascade screening of SDR (SDR subtree). The SDR subtree is attached to a Markov model to reflect the SDR’s follow-up pathway. We did not model cascade screening beyond SDR. All screening protocols shared the same decision tree structure but were populated with different parameter values. Probability parameters in the subtrees vary according to DLCN and genetic risk classification of the proband. Screening cost parameters vary depending on the use of DLCN and/or genetic tests.

The Markov cohort model for disease progression (**Figure A7** in **Appendix 4**) included five health states: 1) No CVD, 2) CVD onset and alive, 3) CVD onset and dead, 4) Post-CVD, and 5) Death, with cycle length of one year. All probands and FDR/ SDR were assumed to be CVD-free. Hence, all patients enter the model through the “No CVD” health state. Each year, it is possible to stay CVD-free, develop CVD or die. The probabilities for developing CVD are stratified by age, FH status, type of medication and length of time (in years) since starting treatment. Once a patient develops CVD, he/she enters the tunnel states “CVD onset and alive” or “CVD onset and dead”, depending on whether the patients survived the CVD event. In the next cycle, patients who survived the event will transit to “Post-CVD” health state, while patients who died from the event will transit to “Death”, an absorbing state. In the following cycles, post-CVD patients may either live with CVD or die.

The Markov model did not set recurrent CVD events as an individual health state, due to lack of data on recurrence rates in Singapore. Instead, we modelled the secondary prevention effect of PCSK9i by assuming that health-related quality of life (HRQoL) loss due to CVD is reduced by 17%, since PCSK9i is found to reduce major cardiovascular events by 17% without significant impact on mortality [16].

The model was built using Excel for Microsoft 365® as this is required by Singapore’s HTA agency.

### 2.9. Measurement and valuation of resources and costs

The material/ manpower/ drug costs during screening and follow-up management before CVD onset were quantified using bottom-up approach based on inputs from FHCARE registry, local clinicians and local hospital databases. Annual additional medical cost living with CVD was estimated based on nationwide claim data during 1990-2019.

### 2.10. Currency, price date, and conversion

Costs were expressed in US dollars as of 2023, except the price of PCSK9i and genetic tests which were as of 2024 as these reflect the most recent prices available to us. Exchange rate for year 2022 was used for cross-currency conversion. Consumer price index in Singapore was used for year-on-year conversion.

### 2.11. Selection, measurement and valuation of outcomes

The health outcome was measured in terms of quality-adjusted life years (QALYs), based on EQ-5D utility scores of Singapore general population and CVD patients from a local study [17]. The final outcomes included incremental cost per person screened (Δcost), incremental QALYs per person screened (ΔQALY), incremental cost-effectiveness ratio (ICER), probability of being cost-effective and probability of being most cost-effective across all screening protocols.

All the outcome measures in the base case analysis were the average values based on two thousand simulations in probabilistic sensitivity analysis (PSA). We adopted probabilistic measures instead of deterministic measures, because nonlinearity of the model leads to systematic and sizable downward bias in deterministic ICER (**Table A4** in **Appendix 6**). Using probabilistic measures in base case analysis is recommended by NICE [18].

### 2.12. Characterizing uncertainty

Standard error (SE) of the point estimates of each parameter were used to populate the probabilistic model. Two-way plot of probabilities of being cost-effective and being most cost-effective across the range of cascade acceptance rates were generated using probabilistic model. Deterministic sensitivity analysis (DSA) was conducted by varying each of 63 independent parameters (or parameter groups) from its base case value across a range of values. This will enable parameters to be identified that were influential to ICER and thus to suggest areas for refining the screening over time. For a fair comparison in DSA, SE and range was set consistently across parameters.

In the base case, we assumed patients start treatment immediately on diagnosis. In reality, it may take some time for patients to get started on and adhere to lipid-lowering treatment. Thus, we conducted an extreme scenario analysis assuming all patients start treatment 5 years after screening.

## 3. Results

### 3.1. Base-case analysis

**Table 1** shows the averaged results from 2000 rounds of parameter simulation for base case scenario where PCSK9i is not considered for treatment and acceptance rates for cascade screening is the realistic values in Singapore: about 50% of probands share the contact of their first-degree relatives (FDR) and about 10% or 50% of FDR contacted agree to be screened, depending on if a pathogenic variant was detected in the proband. All screening protocols are very likely to be cost-effective, with ICERs ranging between $19k-30k/QALY (cost-effectiveness threshold, CET = $41k/QALY) and probabilities of being cost-effective ranging between 85-95%. The most cost-effective protocol is offering NGS to probands with DLCN > 5 (P2), with the smallest ICER ($19k/QALY) and highest probability of being most cost-effective (84%).

### 3.2. One-way DSA

In the most cost-effective protocol (P2), cascade acceptance rate is the top driver of ICER (**Figure 1**). Across all screening protocols (**Figure A8** in **Appendix 6**), cascade acceptance rate is consistently the top driver of ICER. Other drivers of ICER common across multiple protocols include age of proband, reduction in HRQoL with CVD, prevalence of FH among probands, risk of CVD associated with genetic FH, lipid-lowering treatment effect and manpower use during treatment pre-CVD.

**Table 1.**
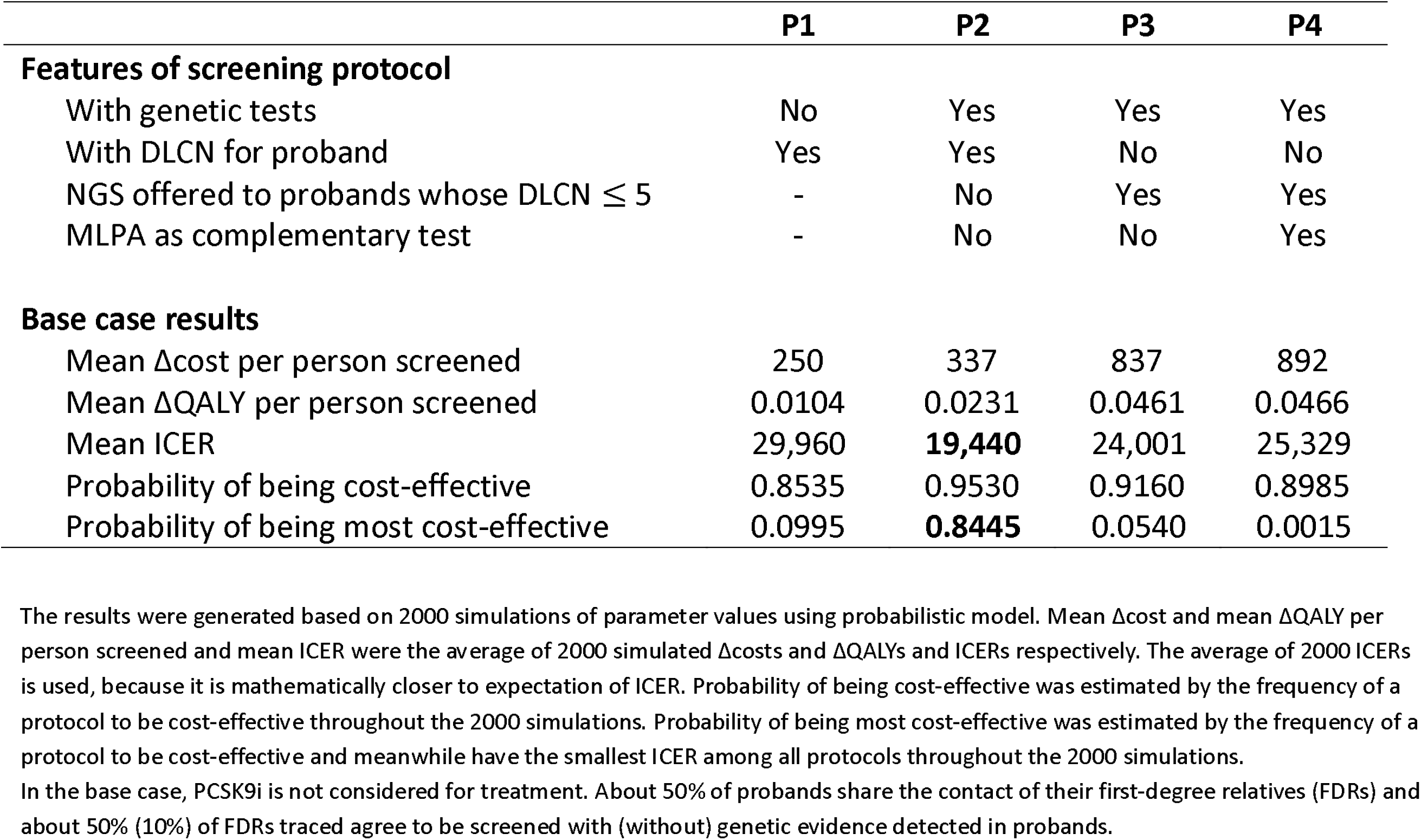
Base case results: Cost-effectiveness of various screening protocols for FH.

**Figure 1.**
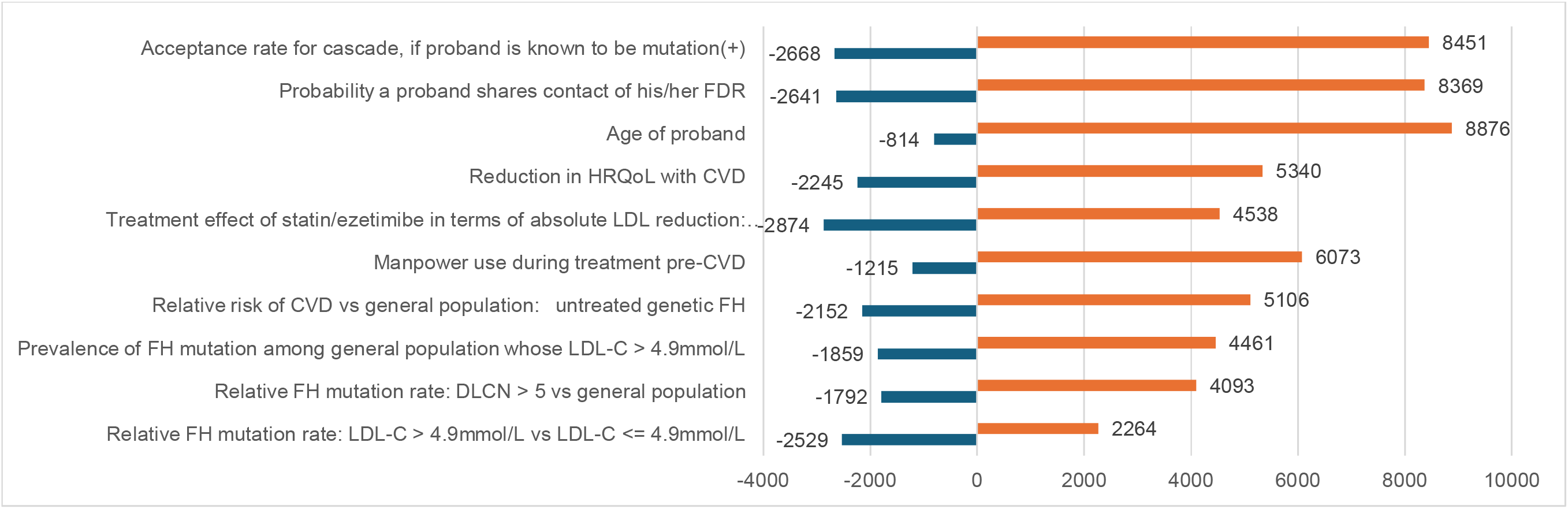
One-way DSA: Top-10 drivers for ICER of Protocol 2 in base case scenario The tornado graph was generated using deterministic model.

### 3.3. Various cascade acceptance rates

**Figure 2** displays the change in the likelihoods for each screening protocol to be cost-effective and most cost-effective, when cascade acceptance rates (including probability for probands to share contact of FDR and probability for traced FDR to agree to be screened with mutation detected in probands) move simultaneously from 0 to 100%, with probability for contacted FDR to agree to be screened kept at a ratio of 1:5 for probands without and with detected pathogenic variants. When both probabilities (proband to provide contact of FDR and FDR to accept screening) are 30% and below, all screening protocols are unlikely to be cost-effective. Across all cascade acceptance rates, Protocol 2 is consistently the most cost-effective.

Most published studies assumed 100% cascade acceptance rates regardless of whether probands have detected pathogenic variants or not. If we make the same assumptions, then ICERs for all protocols would range between $4k-6k/QALY, a difference of about 80% from base case ICERs (Table A2 in Appendix 5). This is the ceiling of cost-effectiveness by improving cascade acceptance rates, although perfect acceptance rates are impossible to achieve.

**Figure 2.**
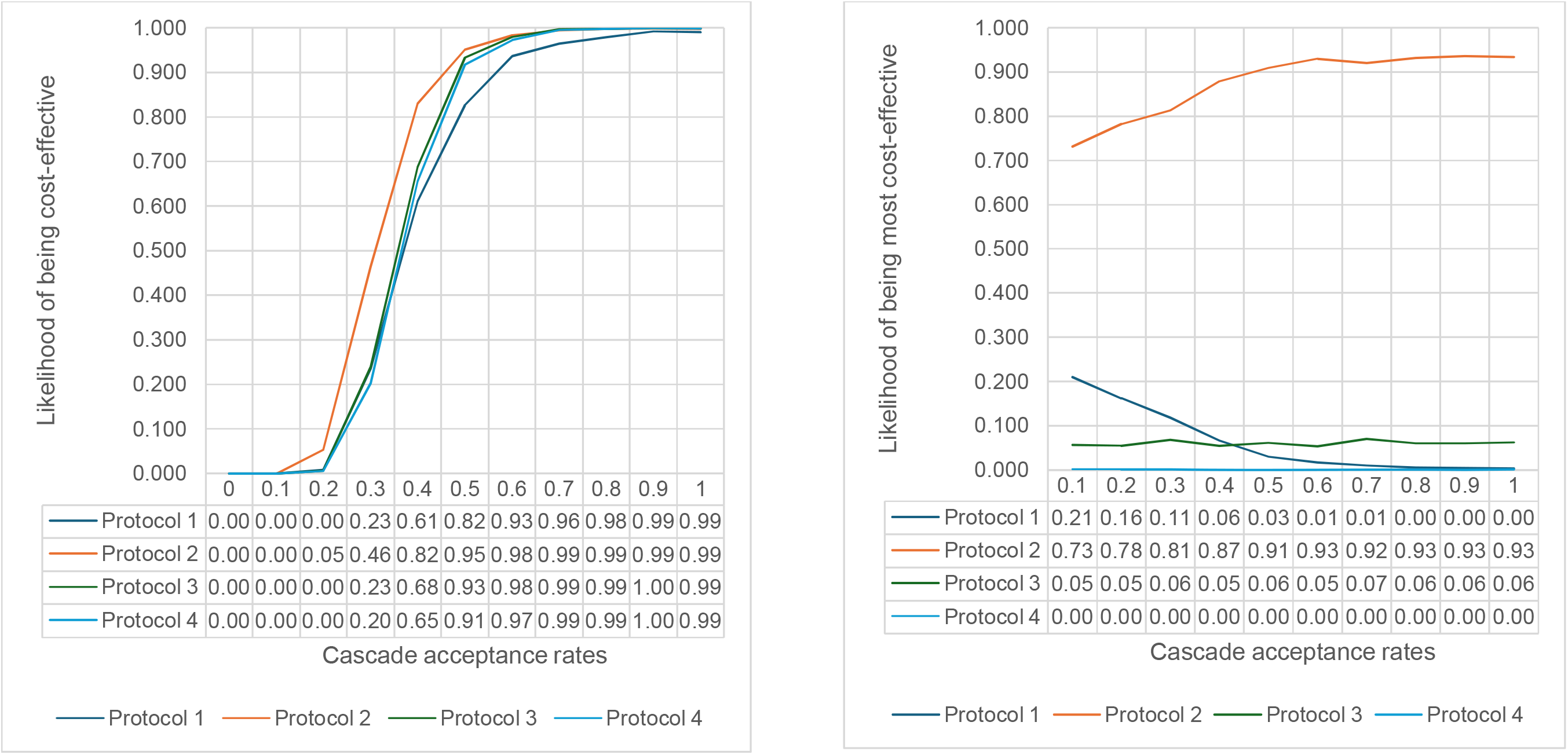
How cascade acceptance rates affect likelihood of being cost-effective and being most cost-effective Cascade acceptance rates (including probability for probands to share contact of FDRs and probability for traced FDRs to agree to be screened with mutation detected in probands) were assumed to move simultaneously from 0 to 100% with step length of 10%. Probability for traced FDRs to agree to be screened without mutation is kept as 1/5 of that with mutation. Given a set of cascade acceptance rates, the probabilities were generated based on 2000 simulations using probabilistic model.

### 3.4. Scenario analysis: start treatment 5 years after screening

If all newly diagnosed patients start treatment 5 years after the screening (**Table 2**), ICERs increase by $10k-21k/QALY, which are sizable compared to the ICER changes in Figure 1. P2 remains to be the most cost-effective protocol. P1 and P2 are still likely to be cost-effective (ICERs < CET = $41k/QALY; 71% to 77% to be cost-effective), while P3 and P4’s cost-effectiveness is uncertain (ICERs > CET = $41k/QALY; 61% to 64% to be cost-effective). Therefore, timeliness of starting treatment is another driver for ICER.

**Table 2.**
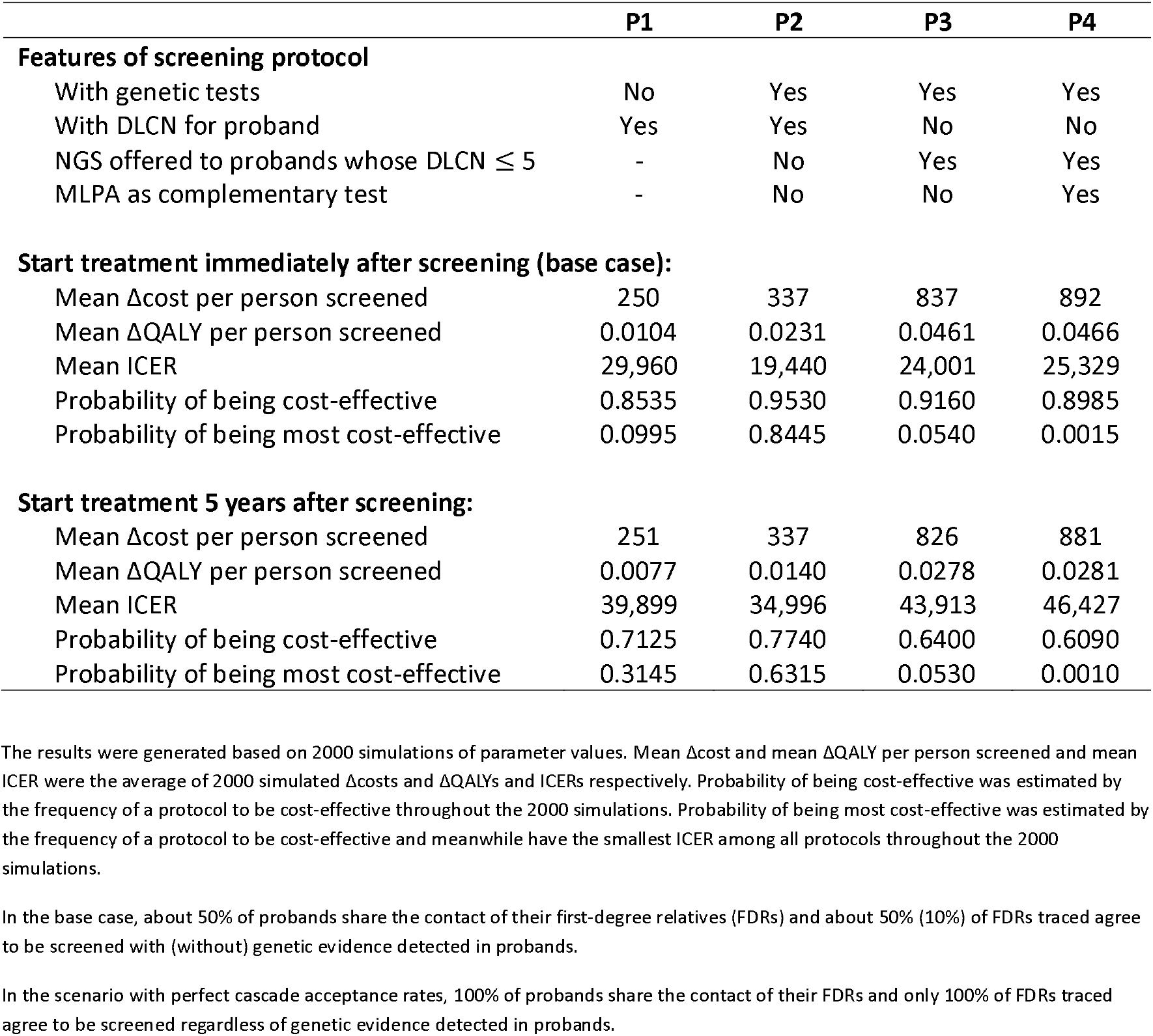
Cost-effectiveness if treatment is delayed for 5 years.

|  | P1 | P2 | P3 | P4 |
| --- | --- | --- | --- | --- |
| **Features of screening protocol** |  |  |  |  |
| With genetic tests | No | Yes | Yes | Yes |
| With DLCN for proband | Yes | Yes | No | No |
| NGS offered to probands whose DLCN $\leq 5$ | - | No | Yes | Yes |
| MLPA as complementary test | - | No | No | Yes |
| **Start treatment immediately after screening (base case):** |  |  |  |  |
| Mean Δcost per person screened | 250 | 337 | 837 | 892 |
| Mean ΔQALY per person screened | 0.0104 | 0.0231 | 0.0461 | 0.0466 |
| Mean ICER | 29,960 | 19,440 | 24,001 | 25,329 |
| Probability of being cost-effective | 0.8535 | 0.9530 | 0.9160 | 0.8985 |
| Probability of being most cost-effective | 0.0995 | 0.8445 | 0.0540 | 0.0015 |
| **Start treatment 5 years after screening:** |  |  |  |  |
| Mean Δcost per person screened | 251 | 337 | 826 | 881 |
| Mean ΔQALY per person screened | 0.0077 | 0.0140 | 0.0278 | 0.0281 |
| Mean ICER | 39,899 | 34,996 | 43,913 | 46,427 |
| Probability of being cost-effective | 0.7125 | 0.7740 | 0.6400 | 0.6090 |
| Probability of being most cost-effective | 0.3145 | 0.6315 | 0.0530 | 0.0010 |

The results were generated based on 2000 simulations of parameter values. Mean Δcost and mean ΔQALY per person screened and mean ICER were the average of 2000 simulated $\Delta$ costs and $\Delta$ QALYs and ICERs respectively. Probability of being cost-effective was estimated by the frequency of a protocol to be cost-effective throughout the 2000 simulations. Probability of being most cost-effective was estimated by the frequency of a protocol to be cost-effective and meanwhile have the smallest ICER among all protocols throughout the 2000 simulations.
In the base case, about 50% of probands share the contact of their first-degree relatives (FDRs) and about 50% (10%) of FDRs traced agree to be screened with (without) genetic evidence detected in probands.
In the scenario with perfect cascade acceptance rates, 100% of probands share the contact of their FDRs and only 100% of FDRs traced agree to be screened regardless of genetic evidence detected in probands.

### 3.5. Scenario analysis: PCSK9i accessible for FH and CVD patients

With accessibility to PCSK9i and realistic cascade acceptance rates (**Table 3**), the most cost-effective protocol is offering NGS to any probands whose LDL-c > 4.9mmol/L without MLPA (P3). All screening protocols remain likely to be cost-effective (73% to 99%). Compared to base case, P1 and P2 are less cost-effective (larger ICER, lower likelihood to be cost-effective), while P3 and P4 are more cost-effective (smaller ICER, higher likelihood to be cost-effective). This is mainly because P1 and P2 provides PCSK9i to clinical FH probands based on DLCN, while P3 and P4 limit PCSK9i access to genetic FH probands and relatives only. Although it is cost-effective to provide PCSK9i for clinical FH, it is not as cost-effective as limiting access to PCSK9i for genetic FH only. This is because genetic FH has more than 3-fold CVD risk compared to clinical FH [19].

**Table 3.** Cost-effectiveness of various screening protocols in the scenario where PCSK9i is accessible for FH and CVD patients.

|  | P1 | P2 | P3 | P4 |
| --- | --- | --- | --- | --- |
| **Features of screening protocol** |  |  |  |  |
| With genetic tests | No | Yes | Yes | Yes |
| With DLCN for proband | Yes | Yes | No | No |
| NGS offered to probands whose DLCN $\leq 5$ | - | No | Yes | Yes |
| MLPA as complementary test | - | No | No | Yes |
| **No access to PCSK9i (base case):** |  |  |  |  |
| Mean Δcost per person screened | 250 | 337 | 837 | 892 |
| Mean ΔQALY per person screened | 0.0104 | 0.0231 | 0.0461 | 0.0466 |
| Mean ICER | 29,960 | 19,440 | 24,001 | 25,329 |
| Probability of being cost-effective | 0.8535 | 0.9530 | 0.9160 | 0.8985 |
| Probability of being most cost-effective | 0.0995 | 0.8445 | 0.0540 | 0.0015 |
| **PCSK9i is accessible for FH and CVD patients:** |  |  |  |  |
| Mean Δcost per person screened | 1,944 | 2,236 | 1,893 | 1,960 |
| Mean ΔQALY per person screened | 0.0624 | 0.0882 | 0.1071 | 0.1082 |
| Mean ICER | 34,045 | 27,851 | **19,843** | 20,389 |
| Probability of being cost-effective | 0.7305 | 0.8595 | 0.9890 | 0.9875 |
| Probability of being most cost-effective | 0.0385 | 0.1420 | **0.8195** | 0.0000 |

FH patients are defined as those with FH mutation, or whose DLCN $> 5$ but with no mutation. About 50% of probands share the contact of their first-degree relatives (FDRs) and about 50% (10%) of FDRs traced agree to be screened with (without) genetic evidence detected in probands. The results were generated based on 2000 simulations using probabilistic model. Mean Δcost and mean ΔQALY per person screened and mean ICER were the average of 2000 simulated $\Delta$ costs and $\Delta$ QALYs and ICERs respectively. Probability of being cost-effective was estimated by the frequency of a protocol to be cost-effective throughout the 2000 simulations. Probability of being most cost-effective was estimated by the frequency of a protocol to be cost-effective and meanwhile have the smallest ICER among all protocols throughout the 2000 simulations.

Analyses in **Section 3.2** to **3.4** were replicated for the scenario with access to PCSK9i (**Table A3** and **Table A4** in **Appendix 5**; **Figure A9** and **A10** in **Appendix 6**). Cascade acceptance rates, timeliness of starting treatment, age of proband, reduction in HRQoL with CVD, prevalence of FH among probands and conventional lipid-lowering treatment effects remain as common drivers for ICER, with cost of PCSK9i being an additional driver (since PCSK9i is a high-cost drug). The most cost-effective screening protocol is P1 with low cascade acceptance rates, but switches to P3 when cascade acceptance rates grow high enough for health gains from relatives to successfully offset costs of genetic tests.

## 4. Discussion

This study aims to evaluate the cost-effectiveness of alternative cascade screening protocols, understand the roles of cascade acceptance rates and high-cost drugs, and look for directions to improve cost-effectiveness of cascade screening for FH.

### 4.1. Is cascade screening for FH cost-effective?

In the Singapore context, all cascade screening protocols for FH are likely to be cost-effective, despite the use of high-cost drug (PCSK9i) and moderate cascade acceptance rates (50% of probands share the contact of their FDR; of these, if a causative variant is identified, 50% agree to be screened, yet in absence of a causal variant, only 10% opt for clinical screening). The generalizability of this finding to other countries and settings will depend on the cost of PCSK9i as well as cascade acceptance rates, among other factors.

### 4.2. What is the most cost-effective cascade screening protocol? Is screening with genetic testing more cost-effective than screening without genetic testing?

In the Singapore context, the most cost-effective screening protocol is to offer NGS to any probands whose LDL-c > 4.9mmol/L without MLPA as complementary test for NGS. The nationwide screening program to be launched in Singapore by mid-2025 is consistent with this strategy [20].

In other contexts, depending on cascade acceptance rates and choice of lipid-lowering medications, the answers may differ. A certain level of cascade acceptance rates may be required for screening with genetic tests to be more cost-effective than screening without genetic tests. Adding MLPA as a complement for NGS may not be as cost-effective as using NGS alone, because the yield from MLPA is not high among NGS-negative probands. NGS combined with bioinformatics calculation can detect most copy number variations (CNVs) now, and the technology will advance over time.

### 4.3. How does cascade acceptance rates affect cost-effectiveness of the screening, and what are policy implications?

Cascade acceptance rates are key drivers of ICER, comprising (i) proband’s acceptance rate to share contacts of his/her relatives and (ii) relatives’ acceptance rate to be screened. The two cascade acceptance rates affect ICER mainly through their product, FDR participation rate, so independently the two acceptance rates have almost the same effect on ICER. Higher acceptance rates for cascade normally improve the cost-effectiveness of the screening. Low cascade acceptance rates may lead to non-cost-effectiveness of cascade screening. Other acceptance rates during the screening, comprising (i) probability that a proband show up for screening and (ii) proband’s acceptance rate for NGS, were not found as drivers of ICER.

Considering the role of cascade acceptance rates, we would recommend countries to (i) use realistic acceptance rates instead of perfect acceptance rates when evaluating cost-effectiveness of the cascade screening in their setting, (ii) identify proband subgroups with better cascade acceptance rates, and (iii) find ways to improve cascade acceptance rates over time. Regarding (ii), for example, we found probands under 26 years old have significantly higher FDR participation rate (40%) than those above 26 years old (12%), though the reason for this is unclear and requires further study. Extremely high LDL-c observed in probands is also found to be associated with high FDR participation rate. Regarding (iii), the cascade screening hesitancy across population could be due to privacy concern and/or lack of knowledge and communication skills when probands reach out to their family members. The Netherlands FH programme achieved a very high FDR participation rate (90%) potentially due to the engagement of specialist nurses to directly contact with relatives and in-home visits for sample collection (for laboratory and genetic testing)[21,22]. Singapore is piloting those methods, together with other methods that may potentially improve cascade acceptance rates including utilization of chatbot to interact with patients more frequently.

### 4.4. Does access to high-cost drugs make the screening not cost-effectiveness?

Access to high-cost drug does not necessarily overturn the cost-effectiveness of cascade screening for FH. The reasons are three-fold: first, high-cost drugs may have higher effectiveness; second, although giving PCSK9i incurs high costs, it saves potential future costs associated with CVD; third, as in the Singapore case, prescribing high-cost drug to proband conditional on screening results generates greater clinical benefits.

### 4.5. How to improve cost-effectiveness of cascade screening for FH?

Based on findings from one-way DSA, efficient and practical ways to improve cost-effectiveness of the screening include raising the acceptance rates for cascade screening, focusing on particular age groups for proband screening and improve screening detection rate among probands (but with limited increase in cost of proband identification), and starting treatment in time post-screening. Since improving these key drivers may incur cost, future studies can examine the cost-effectiveness of practices that improve these key drivers.

This study does have a few limitations. First, since there is no life table for FH patients in Singapore, so to model the impact of follow-up treatment, we estimated CVD risks based on LDL-c changes before and after treatment. Second, we did not model screening beyond SDR of the proband or future generations of FH families. Nevertheless, this may not substantially affect the cost-effectiveness results, given that the observed cascade acceptance rates are low and that future benefits tend to be small because of discounting. For example, the cost-effectiveness only marginally differed when SDR of the proband were excluded from the model (**Table A5** and **A6** in **Appendix 5**). Third, the probabilities of screening without genetic test (which was never implemented in Singapore) were assumed or inferred based on various data sources, which may or may not be the actual case. Fourth, for HRQoL with CVD, we used local estimates that were obtained from myocardial infarction and stroke patients [17], which might have overestimated the QALY loss from other CVD conditions (e.g. angina). Nonetheless, current literature suggests that HRQoL of angina is comparable to the values that we have used in our analyses [23]. Regardless, as HRQoL with CVD is one of the key drivers of ICER, it is worthwhile to obtain more precise information on the HRQoL estimates. Fifth, the health system perspective was adopted as per Singapore’s health technology assessment (HTA) agency’s guidelines [15]. The choice of health system perspective means that indirect costs from lost productivity and premature death were not accounted for, which may be huge with FH as CVD may develop much earlier during economically productive age. Including indirect cost will make the screening look even more cost-effective, but unlikely to change the conclusions in this study.

### 4.6. Conclusion

Cascade screening for FH is cost-effective across most scenarios and assumptions. In the Singapore context, the most cost-effective screening protocol is to offer NGS to any probands whose LDL-c > 4.9mmol/L without MLPA as complementary test for NGS. Cascade acceptance rates are key drivers of cost-effectiveness. Hence, health systems need to look for ways to improve proband’s willingness to share contact of their relatives and relatives’ willingness to be screened. Access to high-cost drugs may not necessarily lead to non-cost-effectiveness of the screening. Other ways to improve cost-effectiveness of the screening include to focus on particular age groups for proband screening, improve screening detection rate among probands (but without increasing cost of proband identification), and start treatment in time post-screening.

## Data Availability

All data produced in the present work are contained in the manuscript

## Ethics Approval

This study was reviewed and approved by the National Health Group Domain Specific Review Board (2020/01407).

## Acknowledgements

The authors would like to thank E Shyong TAI for providing clinical advice for this study. We appreciate support from Stephanus Evan LUKANTA for providing the LDL-c distribution data of the Singapore Population Health Studies - Multi-ethnic Cohort (SPHS-MEC) from the Saw Swee Hock School of Public Health, National University of Singapore. We would also like to show our gratitude to 1) Sock Hoai CHAN, 2) Patrick TAN, 3) Joanne NGEOW, and 4) Saumya S JAMUAR for their support and contribution in providing prevalence statistics of Familial Hypercholesterolemia among various population groups in Singapore.

