## Appendix for "Cost-effectiveness of alternative cascade screening strategies for familial hypercholesterolemia with realistic cascade screening acceptance rates and use of high-cost drugs"

### Appendix 1. Table of Terms

| **Terminology** | **Definition** |
| --- | --- |
| *Cascade screening for FH* | This may refer to either of below depending on the context:   - the screening on the relatives of a proband who has been detected with DLCN>5 (if screened without genetic testing) or causative variants (if screened with genetic testing) - the screening on a proband with diagnostic tools (DLCN and/or genetic tests) and the screening on his/her relatives |
| *Proband screening for FH* | The screening on a proband with diagnostic tools (DLCN and/or genetic tests) |
| *Proband identification for FH* | The process to identify who can be referred as a proband for the screening for FH. Here, it is, among existing patients at primary/secondary care who took an opportunistic lipid test, look for those with LDL-c > 4.9mmol/L. |
| *Probability of probands to share contacts of their FDR* | This includes both probabilities below which share the same value:   - the probability that a proband shares contacts of his/her FDR - the probability that an FDR shares contacts of his/her FDR (SDR of the proband) |
| *Probability of the contacted FDR to accept screening* | This includes both probabilities below which share the same value:   - the probability that a contacted FDR agree to be screened - the probability that a contacted SDR (of the proband) agree to be screened |
| *Cascade acceptance rates* | This includes both probabilities below:   - Probability of probands to share contacts of their FDR - Probability of the contacted FDR to accept screening |
| *FDR participation rate* | This is the product of the two cascade acceptance rates:   - Probability of probands to share contacts of their FDR - Probability of the contacted FDR to accept screening |
| *Proband acceptance rates* | This includes both probabilities below:   - Probability that a proband show up for screening - Probability that a proband accept to be screened by NGS |

Abbreviations:

- *FH*: familial hypercholesterolemia
- *DLCN*: Dutch Lipid Clinical Network criteria
- *FDR/SDR*: First degree relatives/Second degree relatives

### Appendix 2. Dutch Lipid Clinic Network (DLCN) Diagnostic Criteria for Familial Hypercholesterolemia

| **Criteria** | **Points** |
| --- | --- |
| **Family history** |  |
| First-degree relative with known premature* coronary and vascular disease, OR First-degree relative with known LDL-C level above the 95th percentile | 1 |
| First-degree relative with tendinous xanthomata and/or arcus cornealis, OR Children aged less than 18 years with LDL-C level above the 95th percentile | 2 |
| **Clinical history** |  |
| Patient with premature* coronary artery disease | 2 |
| Patient with premature* cerebral or peripheral vascular disease | 1 |
| **Physical examination** |  |
| Tendinous xanthomata | 6 |
| Arcus cornealis prior to age 45 years | 4 |
| **Cholesterol levels mg/dl (mmol/liter)** |  |
| LDL-C >= 330 mg/dL (≥ 8.5) | 8 |
| LDL-C 250 – 329 mg/dL (6.5–8.4) | 5 |
| LDL-C 190 – 249 mg/dL (5.0–6.4) | 3 |
| LDL-C 155 – 189 mg/dL (4.0–4.9) | 1 |
| **Diagnosis (diagnosis is based on the total number of points obtained)** |  |
| Definite Familial Hypercholesterolemia | >8 |
| Probable Familial Hypercholesterolemia | 6 – 8 |
| Possible Familial Hypercholesterolemia | 3 – 5 |
| Unlikely Familial Hypercholesterolemia | <3 |

* Premature = < 55 years in men; < 60 years in women

LDL-C = low density lipoprotein cholesterol; FH, familial hypercholesterolemia.

Adapted from the 2016 ESC/EAS guidelines for the management of dyslipidaemia.

### Appendix 3. Diagrams for Screening Protocols

**Figure A1**. Process diagram of screening **Protocol 1**

…………………

FDR agrees to be screened

Ask proband to share FDR’s contact

DLCN

LDL-c > 4.9mmol/L

DLCN > 5

Opportunistic lipid test for proband

Lipid test

Ask FDR to share SDR’s contact

Refer proband for screening

Proband shows up for screening

Invite FDR for screening

Proband agrees to share contact

LDL-c > 4.9mmol/L

FDR agrees to share contact

Abbreviations:

- *LDL-c*: low-density lipoprotein-cholesterol
- *DLCN*: Dutch Lipid Clinical Network criteria
- *FDR/SDR*: First degree relatives/Second degree relatives

**Figure A2**. Process diagram of screening **Protocol 2**

Abbreviations:

- *LDL-c*: low-density lipoprotein-cholesterol
- *DLCN*: Dutch Lipid Clinical Network criteria
- *NGS*: next generation sequencing
- *FDR/SDR*: First degree relatives/Second degree relatives
- *SS*: Sanger sequencing

…………………

FDR agrees to be screened

NGS

Ask proband to share FDR’s contact

NGS (+)

DLCN

LDL-c > 4.9mmol/L

DLCN > 5

Opportunistic lipid test for proband

SS & Lipid test

Ask FDR to share SDR’s contact

Refer proband for screening

Proband shows up for screening

Invite proband for NGS

Invite FDR for screening

Proband agrees to share contact

SS (+)

Proband agrees to take genetic test

FDR agrees to share contact

**Figure A3**. Process diagram of screening **Protocol 3**

…………………

FDR agrees to be screened

NGS

Ask proband to share FDR’s contact

NGS (+)

LDL-c > 4.9mmol/L

Opportunistic lipid test for proband

SS & Lipid test

Ask FDR to share SDR’s contact

Refer proband for screening

Proband shows up for screening

Invite proband for NGS

Invite FDR for screening

Proband agrees to share contact

SS (+)

Proband agrees to take genetic test

FDR agrees to share contact

Abbreviations:

- *LDL-c*: low-density lipoprotein-cholesterol
- *NGS*: next generation sequencing
- *FDR/SDR*: First degree relatives/Second degree relatives
- *SS*: Sanger sequencing

**Figure A4**. Process diagram of screening **Protocol 4**

…………………

NGS (-)

NGS

MLPA

Ask proband to share FDR’s contact

MLPA (+)

NGS (+)

LDL-c > 4.9mmol/L

Opportunistic lipid test for proband

SS & Lipid test

MLPA & Lipid test

FDR agrees to be screened

Proband identified by MPLA

MLPA (+)

FDR agrees to be screened

Proband identified by NGS

Refer proband for screening

Proband shows up for screening

Invite proband for NGS

Invite FDR for screening

Proband agrees to share contact

SS (+)

Proband agrees to take genetic test

Ask FDR to share SDR’s contact

FDR agrees to share contact

Abbreviations:

- *LDL-c*: low-density lipoprotein-cholesterol
- *NGS*: next generation sequencing
- *FDR/SDR*: First degree relatives/Second degree relatives
- *SS*: Sanger sequencing
- *MLPA*: multiplex ligation-dependent probe amplification

### Appendix 4. Model Structures

**Figure A5**. Main decision tree for proband screening

Probands picked up by opportunistic lipid tests were assumed to be previously undiagnosed with FH and CVD-free. The main tree is attached to a Markov model, that reflects the proband’s follow-up pathway (**Figure A7**), and a subtree, for cascade screening of FDRs (**Figure A6**).

**Genetic FH**: FH patients with causative genetic variant from any of 8 genes.)

**Clinically FH**: DLCN $>$ 5, but without causative genetic variant

**Non-FH**: LDL-C $>$ 4.1mmol/L, but DLCN $\leq$ 5 and without causative genetic variant)

Varied FH group

Patients

No show

Show up for screening

DLCN ≤ 5

Reject NGS

Accept NGS

NGS (-)

MLPA (-)

MLPA (+)

NGS (+)

DLCN > 5

Reject NGS

without mutation

with mutation

Accept NGS

NGS (-)

MLPA (-)

MLPA (+)

NGS (+)

Abbreviations:

- *FH*: familial hypercholesterolemia
- *LDL-c*: low-density lipoprotein-cholesterol
- *NGS*: next generation sequencing
- *MLPA*: multiplex ligation-dependent probe amplification
- *DLCN*: Dutch Lipid Clinical Network criteria

**Figure A6**. Subtree for FDR screening (same structure for SDR screening)

The FDR subtree is attached to a Markov model (**Figure A7**) that reflects the FDR’s follow-up pathway and another subtree for cascade screening of SDRs. The SDR subtree is attached to a Markov model to reflect the SDR’s follow-up pathway. The branches of the subtree depend on results from lipid panel and genetic tests, whether the FDR/ SDR was already on lipid-lowering treatment or not, and the DLCN score of the relative. Note that collecting information on the DLCN score of FDR/ SDR, which determines their potential gain from diagnosis and treatment, is currently not a standard practice. The DLCN information of FDR/SDR was collected in FHCARE for research purposes.

**Genetic FH**: FH patients with causative genetic variant from any of 8 genes.)

**Clinically FH**: DLCN $>$ 5, but without causative genetic variant

**Non-FH**: LDL-c $>$ 4.1mmol/L, but DLCN $\leq$ 5 and without causative genetic variant)

Varied FH group

Ask proband for contact of FDR

Contact not shared

Contact shared

FDR reject screening

FDR accept screening

Without mutation

On treatment

Not on treatment

With mutation

On treatment

Not on treatment

Without mutation

On treatment

Not on treatment

With mutation

On treatment

Not on treatment

Abbreviations:

- *FDR/SDR*: First degree relatives/Second degree relatives
- *FH*: familial hypercholesterolemia
- *LDL-c*: low-density lipoprotein-cholesterol
- *DLCN*: Dutch Lipid Clinical Network criteria

**Figure A7**. Markov Model Diagram for Follow-up Treatment and Cardiovascular Disease Progression

All probands and FDRs/ SDRs were assumed to be CVD-free for the purpose of simplifying. Hence, all patients enter the model through the “No CVD” health state.

**CVD onset**

**and alive**

**CVD onset**

**and dead**

Abbreviations:

- *CVD:* Cardiovascular Disease

### Appendix 5. Additional Result Tables

**Table A1**. **Deterministic ICER** (with systematic downward bias) and probabilistic ICER

|  | **P1** | **P2** | **P3** | **P4** |
| --- | --- | --- | --- | --- |
| **Features of screening protocol** |  |  |  |  |
| With genetic tests | No | Yes | Yes | Yes |
| With DLCN for proband | Yes | Yes | No | No |
| NGS offered to probands whose DLCN $\leq$ 5 | - | No | Yes | Yes |
| MLPA as complementary test | - | No | No | Yes |
| **Base case:** | | | | |
| Deterministic ICER | 3,445 | 2,183 | 2,763 | 2,904 |
| Probabilistic Mean ICER *(base case)* | 5,450 | 4,654 | 5,300 | 5,505 |
| **Scenario with access to PCSK9i:** |  |  |  |  |
| Deterministic ICER | 23,850 | 18,841 | 12,163 | 12,531 |
| Probabilistic Mean ICER *(base case)* | 34,045 | 27,851 | 19,843 | 20,389 |

The deterministic ICER was generated based on the mean value of each parameter.

The probabilistic ICER was the mean value of 2000 ICERs based on 2000 simulations of parameter values.

**Table A2**. Cost-effectiveness with **perfect cascade acceptance rates** (no access to PCSK9i)

|  | **P1** | **P2** | **P3** | **P4** |
| --- | --- | --- | --- | --- |
| **Features of screening protocol** |  |  |  |  |
| With genetic tests | No | Yes | Yes | Yes |
| With DLCN for proband | Yes | Yes | No | No |
| NGS offered to probands whose DLCN $\leq$ 5 | - | No | Yes | Yes |
| MLPA as complementary test | - | No | No | Yes |
| **Base case cascade acceptance rates:** | | | | |
| Mean Δcost per person screened | 250 | 337 | 837 | 892 |
| Mean ΔQALY per person screened | 0.0104 | 0.0231 | 0.0461 | 0.0466 |
| Mean ICER | 29,960 | 19,440 | 24,001 | 25,329 |
| Probability of being cost-effective | 0.8535 | 0.9530 | 0.9160 | 0.8985 |
| Probability of being most cost-effective | 0.0995 | 0.8445 | 0.0540 | 0.0015 |
| **Perfect cascade acceptance rates:** | | | | |
| Mean Δcost per person screened | 756 | 602 | 1,285 | 1,345 |
| Mean ΔQALY per person screened | 0.1501 | 0.1571 | 0.2959 | 0.2989 |
| Mean ICER | 5,450 | 4,654 | 5,300 | 5,505 |
| Probability of being cost-effective | 0.9995 | 0.9990 | 0.9995 | 0.9995 |
| Probability of being most cost-effective | 0.2420 | 0.7210 | 0.0370 | 0.0010 |

The results were generated based on 2000 simulations of parameter values. Mean Δcost and mean ΔQALY per person screened and mean ICER were the average of 2000 simulated Δcosts and ΔQALYs and ICERs respectively. Probability of being cost-effective was estimated by the frequency of a protocol to be cost-effective throughout the 2000 simulations. Probability of being most cost-effective was estimated by the frequency of a protocol to be cost-effective and meanwhile have the smallest ICER among all protocols throughout the 2000 simulations.

In the base case, about 50% of probands share the contact of their first-degree relatives (FDRs) and about 50% (10%) of FDRs traced agree to be screened with (without) genetic evidence detected in probands.

In the scenario with perfect cascade acceptance rates, 100% of probands share the contact of their FDRs and only 100% of FDRs traced agree to be screened regardless of genetic evidence detected in probands.

**Table A3**. Cost-effectiveness with **perfect cascade acceptance rates** (scenario with access to PCSK9i)

|  | **P1** | **P2** | **P3** | **P4** |
| --- | --- | --- | --- | --- |
| **Features of screening protocol** |  |  |  |  |
| With genetic tests | No | Yes | Yes | Yes |
| With DLCN for proband | Yes | Yes | No | No |
| NGS offered to probands whose DLCN $\leq$ 5 | - | No | Yes | Yes |
| MLPA as complementary test | - | No | No | Yes |
| **Base case cascade acceptance rates:** | | | | |
| Mean Δcost per person screened | 1,944 | 2,236 | 1,893 | 1,960 |
| Mean ΔQALY per person screened | 0.0624 | 0.0882 | 0.1071 | 0.1082 |
| Mean ICER | 34,045 | 27,851 | 19,843 | 20,389 |
| Probability of being cost-effective | 0.7305 | 0.8595 | 0.9890 | 0.9875 |
| Probability of being most cost-effective | 0.0385 | 0.1420 | 0.8195 | 0.0000 |
| **Perfect cascade acceptance rates:** | | | | |
| Mean Δcost per person screened | 1,212 | 3,182 | 3,729 | 3,815 |
| Mean ΔQALY per person screened | 0.1689 | 0.2744 | 0.4571 | 0.4618 |
| Mean ICER | 7,864 | 12,831 | 8,849 | 8,976 |
| Probability of being cost-effective | 0.9995 | 0.9995 | 0.9995 | 0.9995 |
| Probability of being most cost-effective | 0.6525 | 0.0010 | 0.3455 | 0.0005 |

The results were generated based on 2000 simulations of parameter values. Mean Δcost and mean ΔQALY per person screened and mean ICER were the average of 2000 simulated Δcosts and ΔQALYs and ICERs respectively. Probability of being cost-effective was estimated by the frequency of a protocol to be cost-effective throughout the 2000 simulations. Probability of being most cost-effective was estimated by the frequency of a protocol to be cost-effective and meanwhile have the smallest ICER among all protocols throughout the 2000 simulations.

In the base case, about 50% of probands share the contact of their first-degree relatives (FDRs) and about 50% (10%) of FDRs traced agree to be screened with (without) genetic evidence detected in probands.

In the scenario with perfect cascade acceptance rates, 100% of probands share the contact of their FDRs and only 100% of FDRs traced agree to be screened regardless of genetic evidence detected in probands.

**Table A4**. Cost-effectiveness with **treatment delayed for 5 years** (scenario with access to PCSK9i)

|  | **P1** | **P2** | **P3** | **P4** |
| --- | --- | --- | --- | --- |
| **Features of screening protocol** |  |  |  |  |
| With genetic tests | No | Yes | Yes | Yes |
| With DLCN for proband | Yes | Yes | No | No |
| NGS offered to probands whose DLCN $\leq$ 5 | - | No | Yes | Yes |
| MLPA as complementary test | - | No | No | Yes |
| **Start treatment immediately after screening (base case):** | | | | |
| Mean Δcost per person screened | 1,944 | 2,236 | 1,893 | 1,960 |
| Mean ΔQALY per person screened | 0.0624 | 0.0882 | 0.1071 | 0.1082 |
| Mean ICER | 34,045 | 27,851 | 19,843 | 20,389 |
| Probability of being cost-effective | 0.7305 | 0.8595 | 0.9890 | 0.9875 |
| Probability of being most cost-effective | 0.0385 | 0.1420 | 0.8195 | 0.0000 |
| **Start treatment 5 years after screening:** | | | | |
| Mean Δcost per person screened | 1,393 | 1,587 | 1,406 | 1,467 |
| Mean ΔQALY per person screened | 0.0444 | 0.0577 | 0.0630 | 0.0637 |
| Mean ICER | 32,776 | 30,295 | 27,986 | 29,097 |
| Probability of being cost-effective | 0.7660 | 0.8205 | 0.9055 | 0.8925 |
| Probability of being most cost-effective | 0.1920 | 0.1640 | 0.6425 | 0.0015 |

The results were generated based on 2000 simulations of parameter values. Mean Δcost and mean ΔQALY per person screened and mean ICER were the average of 2000 simulated Δcosts and ΔQALYs and ICERs respectively. Probability of being cost-effective was estimated by the frequency of a protocol to be cost-effective throughout the 2000 simulations. Probability of being most cost-effective was estimated by the frequency of a protocol to be cost-effective and meanwhile have the smallest ICER among all protocols throughout the 2000 simulations.

In the base case, about 50% of probands share the contact of their first-degree relatives (FDRs) and about 50% (10%) of FDRs traced agree to be screened with (without) genetic evidence detected in probands.

In the scenario with perfect cascade acceptance rates, 100% of probands share the contact of their FDRs and only 100% of FDRs traced agree to be screened regardless of genetic evidence detected in probands.

**Table A5**. Cost-effectiveness with **only FDRs of probands screened** (no access to PCSK9i)

|  | **P1** | **P2** | **P3** | **P4** |
| --- | --- | --- | --- | --- |
| **Features of screening protocol** |  |  |  |  |
| With genetic tests | No | Yes | Yes | Yes |
| With DLCN for proband | Yes | Yes | No | No |
| NGS offered to probands whose DLCN $\leq$ 5 | - | No | Yes | Yes |
| MLPA as complementary test | - | No | No | Yes |
| **Screening both FDRs and SDRs of the proband (base case):** | | | | |
| Mean Δcost per person screened | 250 | 337 | 837 | 892 |
| Mean ΔQALY per person screened | 0.0104 | 0.0231 | 0.0461 | 0.0466 |
| Mean ICER | 29,960 | 19,440 | 24,001 | 25,329 |
| Probability of being cost-effective | 0.8535 | 0.9530 | 0.9160 | 0.8985 |
| Probability of being most cost-effective | 0.0995 | 0.8445 | 0.0540 | 0.0015 |
| **Screening only FDRs of the proband:** | | | | |
| Mean Δcost per person screened | 242 | 319 | 808 | 863 |
| Mean ΔQALY per person screened | 0.0102 | 0.0180 | 0.0358 | 0.0361 |
| Mean ICER | 28,773 | 23,195 | 29,032 | 30,687 |
| Probability of being cost-effective | 0.8520 | 0.9140 | 0.8475 | 0.8215 |
| Probability of being most cost-effective | 0.2320 | 0.7075 | 0.0600 | 0.0000 |

The results were generated based on 2000 simulations of parameter values. Mean Δcost and mean ΔQALY per person screened and mean ICER were the average of 2000 simulated Δcosts and ΔQALYs and ICERs respectively. Probability of being cost-effective was estimated by the frequency of a protocol to be cost-effective throughout the 2000 simulations. Probability of being most cost-effective was estimated by the frequency of a protocol to be cost-effective and meanwhile have the smallest ICER among all protocols throughout the 2000 simulations.

In the base case, about 50% of probands share the contact of their first-degree relatives (FDRs) and about 50% (10%) of FDRs traced agree to be screened with (without) genetic evidence detected in probands.

In the scenario with perfect cascade acceptance rates, 100% of probands share the contact of their FDRs and only 100% of FDRs traced agree to be screened regardless of genetic evidence detected in probands.

**Table A6**. Cost-effectiveness with **only FDRs of probands screened** (scenario with access to PCSK9i)

|  | **P1** | **P2** | **P3** | **P4** |
| --- | --- | --- | --- | --- |
| **Features of screening protocol** |  |  |  |  |
| With genetic tests | No | Yes | Yes | Yes |
| With DLCN for proband | Yes | Yes | No | No |
| NGS offered to probands whose DLCN $\leq$ 5 | - | No | Yes | Yes |
| MLPA as complementary test | - | No | No | Yes |
| **Screening both FDRs and SDRs of the proband (base case):** | | | | |
| Mean Δcost per person screened | 1,944 | 2,236 | 1,893 | 1,960 |
| Mean ΔQALY per person screened | 0.0624 | 0.0882 | 0.1071 | 0.1082 |
| Mean ICER | 34,045 | 27,851 | 19,843 | 20,389 |
| Probability of being cost-effective | 0.7305 | 0.8595 | 0.9890 | 0.9875 |
| Probability of being most cost-effective | 0.0385 | 0.1420 | 0.8195 | 0.0000 |
| **Screening only FDRs of the proband:** | | | | |
| Mean Δcost per person screened | 1,921 | 2,164 | 1,780 | 1,845 |
| Mean ΔQALY per person screened | 0.0626 | 0.0807 | 0.0909 | 0.0919 |
| Mean ICER | 33,770 | 29,488 | 21,759 | 22,392 |
| Probability of being cost-effective | 0.7310 | 0.8200 | 0.9770 | 0.9755 |
| Probability of being most cost-effective | 0.0770 | 0.1290 | 0.7935 | 0.0005 |

The results were generated based on 2000 simulations of parameter values. Mean Δcost and mean ΔQALY per person screened and mean ICER were the average of 2000 simulated Δcosts and ΔQALYs and ICERs respectively. Probability of being cost-effective was estimated by the frequency of a protocol to be cost-effective throughout the 2000 simulations. Probability of being most cost-effective was estimated by the frequency of a protocol to be cost-effective and meanwhile have the smallest ICER among all protocols throughout the 2000 simulations.

In the base case, about 50% of probands share the contact of their first-degree relatives (FDRs) and about 50% (10%) of FDRs traced agree to be screened with (without) genetic evidence detected in probands.

In the scenario with perfect cascade acceptance rates, 100% of probands share the contact of their FDRs and only 100% of FDRs traced agree to be screened regardless of genetic evidence detected in probands.

### Appendix 6. Additional Result Figures

**Figure A8**. One-way DSA of all screening protocols under base case scenario (PCSK9i not considered for treatment)

Protocol 1

|  | Column2 | Column3 |
| --- | --- | --- |
| Relative risk of CVD vs general population: untreated genetic FH | -2180 | 3604 |
| Treatment effect of statin/ezetimibe in terms of absolute LDL reduction: genetic FH | -2794 | 3326 |
| Probability a proband show up for screening after referral | -1730 | 5766 |
| Age of proband | 0 | 8823 |
| Prevalence of DLCN > 5 among primary care patients whose LDL-C > 4.9mmol/L | -6296 | 4164 |
| Reduction in HRQoL with CVD | -3430 | 7333 |
| Probability a proband shares contact of his/her FDR | -3294 | 9895 |
| Acceptance rate for cascade, if proband is known to be mutation(+) | -3464 | 10402 |
| Manpower use during treatment pre-CVD | -2421 | 12104 |
| Relative cascade acceptance rate: with genetic evidence vs without genetic evidence | -8011 | 9863 |

Protocol 2

|  | Column2 | Column3 |
| --- | --- | --- |
| Relative FH mutation rate: LDL-C > 4.9mmol/L vs LDL-C <= 4.9mmol/L | -2529 | 2264 |
| Relative FH mutation rate: DLCN > 5 vs general population | -1792 | 4093 |
| Prevalence of FH mutation among general population whose LDL-C > 4.9mmol/L | -1859 | 4461 |
| Relative risk of CVD vs general population: untreated genetic FH | -2152 | 5106 |
| Manpower use during treatment pre-CVD | -1215 | 6073 |
| Treatment effect of statin/ezetimibe in terms of absolute LDL reduction: genetic FH | -2874 | 4538 |
| Reduction in HRQoL with CVD | -2245 | 5340 |
| Age of proband | -814 | 8876 |
| Probability a proband shares contact of his/her FDR | -2641 | 8369 |
| Acceptance rate for cascade, if proband is known to be mutation(+) | -2668 | 8451 |

Protocol 3

|  | Column2 | Column3 |
| --- | --- | --- |
| Manpower use during treatment pre-CVD | -1010 | 5052 |
| Prevalence of DLCN > 5 among primary care patients whose LDL-C > 4.9mmol/L | -4902 | 1544 |
| Relative FH mutation rate: LDL-C > 4.9mmol/L vs LDL-C <= 4.9mmol/L | -4148 | 3713 |
| Treatment effect of statin/ezetimibe in terms of absolute LDL reduction: genetic FH | -3997 | 6249 |
| Prevalence of FH mutation among general population whose LDL-C > 4.9mmol/L | -3048 | 7315 |
| Relative risk of CVD vs general population: untreated genetic FH | -3439 | 7749 |
| Reduction in HRQoL with CVD | -3427 | 8151 |
| Age of proband | -1464 | 14191 |
| Probability a proband shares contact of his/her FDR | -4305 | 13623 |
| Acceptance rate for cascade, if proband is known to be mutation(+) | -4332 | 13705 |

Protocol 4

|  | Column2 | Column3 |
| --- | --- | --- |
| Manpower use during treatment pre-CVD | -1004 | 5018 |
| Prevalence of DLCN > 5 among primary care patients whose LDL-C > 4.9mmol/L | -5312 | 1674 |
| Relative FH mutation rate: LDL-C > 4.9mmol/L vs LDL-C <= 4.9mmol/L | -4495 | 4024 |
| Treatment effect of statin/ezetimibe in terms of absolute LDL reduction: genetic FH | -4223 | 6594 |
| Prevalence of FH mutation among general population whose LDL-C > 4.9mmol/L | -3304 | 7927 |
| Relative risk of CVD vs general population: untreated genetic FH | -3698 | 8282 |
| Reduction in HRQoL with CVD | -3665 | 8717 |
| Age of proband | -1595 | 15209 |
| Probability a proband shares contact of his/her FDR | -4623 | 14617 |
| Acceptance rate for cascade, if proband is known to be mutation(+) | -4650 | 14699 |

The tornado graphs were generated using deterministic model.

**Figure A9**. One-way DSA of all screening protocols under scenario where PCSK9i is accessible for FH and CVD patients

Protocol 1

|  | Column2 | Column3 |
| --- | --- | --- |
| Treatment effect in terms of relative LDL reduction: PCSK9i primary prevention | -357 | 3746 |
| Relative risk of CVD vs general population: untreated clinical FH | -2633 | 2976 |
| Relative FH mutation rate: DLCN > 5 vs general population | -3062 | 3235 |
| Prevalence of FH mutation among general population whose LDL-C > 4.9mmol/L | -3234 | 3443 |
| Relative FH mutation rate: LDL-C > 4.9mmol/L vs LDL-C <= 4.9mmol/L | -5073 | 2163 |
| Annual cost of PCSK9i | -11830 | 0 |
| Relative cascade acceptance rate: with genetic evidence vs without genetic evidence | -10016 | 2041 |
| Reduction in HRQoL with CVD | -5473 | 10116 |
| Treatment effect of statin/ezetimibe in terms of absolute LDL reduction: clinical FH | -7985 | 10998 |
| Age of proband | -10566 | 37890 |

Protocol 2

|  | Column2 | Column3 |
| --- | --- | --- |
| Acceptance rate for NGS by proband | -1221 | 3423 |
| Probability a proband shares contact of his/her FDR | -3240 | 4518 |
| Acceptance rate for cascade, if proband is known to be mutation(+) | -3250 | 4530 |
| Relative FH mutation rate: LDL-C > 4.9mmol/L vs LDL-C <= 4.9mmol/L | -5433 | 3167 |
| Relative FH mutation rate: DLCN > 5 vs general population | -3555 | 5052 |
| Prevalence of FH mutation among general population whose LDL-C > 4.9mmol/L | -3714 | 5379 |
| Annual cost of PCSK9i | -9239 | 0 |
| Treatment effect of statin/ezetimibe in terms of absolute LDL reduction: clinical FH | -5087 | 5842 |
| Reduction in HRQoL with CVD | -4632 | 9111 |
| Age of proband | -5606 | 22691 |

Protocol 3

|  | Column2 | Column3 |
| --- | --- | --- |
| Treatment effect of statin/ezetimibe in terms of absolute LDL reduction: genetic FH | -1014 | 1816 |
| Prevalence of DLCN > 5 among primary care patients whose LDL-C > 4.9mmol/L | -2195 | 691 |
| Relative FH mutation rate: LDL-C > 4.9mmol/L vs LDL-C <= 4.9mmol/L | -1857 | 1662 |
| Additional yearly medical cost living with CVD (vs no CVD including dyslipidemia) | -2987 | 597 |
| Annual cost of PCSK9i | -4220 | 0 |
| Prevalence of FH mutation among general population whose LDL-C > 4.9mmol/L | -1365 | 3275 |
| Probability a proband shares contact of his/her FDR | -2476 | 4952 |
| Acceptance rate for cascade, if proband is known to be mutation(+) | -2489 | 4978 |
| Age of proband | 0 | 9712 |
| Reduction in HRQoL with CVD | -3274 | 7092 |

Protocol 4

|  | Column2 | Column3 |
| --- | --- | --- |
| Treatment effect of statin/ezetimibe in terms of absolute LDL reduction: genetic FH | -1062 | 1889 |
| Prevalence of DLCN > 5 among primary care patients whose LDL-C > 4.9mmol/L | -2378 | 749 |
| Additional yearly medical cost living with CVD (vs no CVD including dyslipidemia) | -2987 | 597 |
| Relative FH mutation rate: LDL-C > 4.9mmol/L vs LDL-C <= 4.9mmol/L | -2012 | 1802 |
| Annual cost of PCSK9i | -4220 | 0 |
| Prevalence of FH mutation among general population whose LDL-C > 4.9mmol/L | -1479 | 3549 |
| Probability a proband shares contact of his/her FDR | -2586 | 5165 |
| Acceptance rate for cascade, if proband is known to be mutation(+) | -2599 | 5192 |
| Age of proband | 0 | 9781 |
| Reduction in HRQoL with CVD | -3373 | 7306 |

The tornado graphs were generated using deterministic model.

**Figure A10**. Scenario with access to PCSK9i: How cascade acceptance rates affect likelihood of being cost-effective and being most cost-effective

Likelihood of being cost-effective

| Cascade acceptance rates | Protocol 1 | Protocol 2 | Protocol 3 | Protocol 4 |
| --- | --- | --- | --- | --- |
| 0 | 0.6175 | 0.5895 | 0.4925 | 0.454 |
| 0.1 | 0.6245 | 0.604 | 0.563 | 0.5265 |
| 0.2 | 0.6405 | 0.6475 | 0.694 | 0.669 |
| 0.3 | 0.6625 | 0.696 | 0.868 | 0.8515 |
| 0.4 | 0.6965 | 0.774 | 0.957 | 0.9505 |
| 0.5 | 0.705 | 0.8415 | 0.986 | 0.984 |
| 0.6 | 0.7485 | 0.93 | 0.997 | 0.996 |
| 0.7 | 0.798 | 0.965 | 0.9965 | 0.996 |
| 0.8 | 0.8305 | 0.989 | 0.999 | 0.999 |
| 0.9 | 0.866 | 0.997 | 1 | 1 |
| 1 | 0.9075 | 0.9995 | 1 | 1 |

Likelihood of being most cost-effective

| Cascade acceptance rates | Protocol 1 | Protocol 2 | Protocol 3 | Protocol 4 |
| --- | --- | --- | --- | --- |
| 0 | 0.558 | 0 | 0.4235 | 0.0005 |
| 0.1 | 0.5225 | 0.0065 | 0.471 | 0 |
| 0.2 | 0.369 | 0.064 | 0.566 | 0.0005 |
| 0.3 | 0.176 | 0.1735 | 0.6495 | 0.001 |
| 0.4 | 0.067 | 0.184 | 0.7485 | 0 |
| 0.5 | 0.0235 | 0.1555 | 0.82 | 0 |
| 0.6 | 0.0085 | 0.113 | 0.878 | 0.0005 |
| 0.7 | 0.0055 | 0.0835 | 0.909 | 0.002 |
| 0.8 | 0.004 | 0.0585 | 0.9365 | 0.001 |
| 0.9 | 0.002 | 0.0495 | 0.948 | 0.0005 |
| 1 | 0.0055 | 0.032 | 0.9615 | 0.001 |

Cascade acceptance rates (including probability for probands to share contact of FDRs and probability for traced FDRs to agree to be screened with mutation detected in probands) were assumed to move from 0 to 100% with step length of 10%. Probability for traced FDRs to agree to be screened without mutation is kept as 1/5 of that with mutation.

Given a set of cascade acceptance rates, the probabilities were generated based on 2000 simulations using probabilistic model.
